# What are the experiences of children and families who use a robotic walker in their home environment? A qualitative study

**DOI:** 10.64898/2026.02.11.26346097

**Authors:** Jessical L. Youngblood, Alicia J. Hilderley, Elizabeth G. Condliffe

## Abstract

**Purpose:** Robotic walkers are a new and novel technology with growing evidence of benefits for children living with mobility impairments. However, little is known about how using these devices at home impacts families. This study aims to explore parents’ perceptions of home-based robotic walking and the impacts on their family and their child living with a mobility impairment.

**Materials and Methods:** Qualitative interviews were conducted with seven parents who have a child who used a robotic walker in their home for at least six months. Thematic analysis was used to analyze all interviews. Themes were then mapped to the F-words for child development.

**Results:** Using a robotic walker at home led to family bonding and created new ways for parents and siblings to interact with the child living with a mobility impairment. Many children enjoyed using the robotic walker. This, combined with being able to direct its use in their own environments, contributed to less parental stress than was associated with other rehabilitation interventions. However, some parents discussed an increase in parental stress due to certain logistical aspects, getting their child in and out and transporting the robotic walker. Finally, parents discussed that obtaining the device was a financial burden for them.

**Conclusion:** Robotic walking in the home environment impacts family relationships and parental stress. Understanding families’ experiences can inform decision-making by families and practitioners around the appropriateness of robotic walker use for a child living with a disability.

## Introduction

Having a child with a mobility impairment can add stress on the family.^1^ One of the main stressors is attending regular clinical appointments for different types of therapies.^2^ Home-based therapies can be advantageous as they may decrease parental stress experienced by parents and may lead to family bonding and increased satisfaction for parents and children.^3,4^ Robotic walking is a new and emerging technology that is typically used in a clinical setting.^5^ However, there is a robotic walker that allows robotic walking to be done at home. There is limited knowledge examining how robotic interventions delivered at home may impact families. This study will focus on how using a robotic walker at home impacts families.

## Theoretical Framework

### The F-words of childhood disability

The conceptual framework of this study is informed by the F-words of child-onset disability framework. ^6^ The F-words are composed of six words that impact every child’s life, however, these areas are typically overlooked in child-onset disability. For a long time, disability was viewed through the medical model lens, where the focus was mainly on ‘fixing’ the individual.^6^ This led to many interventions and treatments focusing on addressing the signs and symptoms of disabilities. ^6,7^ To move beyond this traditional thinking of ‘fixing’ a disability, the World Health Organization created the International Classification of Functioning, Disability, and Health (ICF) framework.^6^ The ICF created a more holistic view of disability that moved beyond impairments and considered the disability in association with personal and environmental factors.^6–8^ However, the ICF model is designed to address many aspects of life experienced in adults with a disability and is not as transferable to childhood disability. ^6,9^ The F-words for childhood disability address this limitation. The F-words are embedded within the ICF to show the interconnectedness of these concepts in the context of childhood disability.^6^

The F-Words are comprised of six words that impact many aspects of children with disabilities’ lives and well-being: Function, Family, Fitness, Fun, Friend, Future. Function is linked to the ICF domain ‘activity’ and focuses on what people do, specifically their ability to perform activities of daily living.^6^ Family is linked to the ICF domain ‘environmental factors’, as family is an essential environment and the main social/personal context for most children with disabilities. Fitness is linked to the ICF domain ‘body structure and function’, as this needs to be emphasized in interventions and health promotion contexts, as children with disabilities tend to have lower levels of fitness. Fun is linked to both the ‘participation’ and ‘personal factor’ ICF domains. Therefore, it focuses on involvement in life which is often overlooked in many clinical and intervention programs. Friends is also linked to the ‘participation’ and ‘personal factor’ domains and is included in this framework to create more meaningful connections for children with disabilities. Finally, Future expands across the entire framework as it is an essential part of development, as researchers and practitioners need to focus more on the future of children living with a disability.^6,8^ This framework is also important to use in research and intervention studies, as it provides researchers and practitioners with concrete examples of how a treatment or technology can impact not only patients/participants, but also their families.^9^ In the context of this present study, the F-words can be used to understand how robotic walking impacts many aspects of the child’s life and family, though more context is needed to understand the full impact on the family.

## Background

Having a child who is unable to walk impacts multiple aspects of family life and can increase parental burden, leading to stress on the parents.^2^ Parents caring for children with a disability often experience stress and isolation due to a lack of community support and difficulties dealing with the healthcare system.^1^ A meta-analytic study with families of young children showed that interventions grounded in family systems theory that provide support and build capacity can improve self-efficacy and interactions among family members, thereby enhancing parent and child well-being.^10^ However, very few of the limited number of evidence-based interventions for individuals with severe mobility impairments are designed for use at home and in the community.

Robotic walking has been shown to have broad positive impacts for children with mobility impairments. Currently, most robotic walking is done in a clinical setting, thus putting stress on the healthcare system.^3,5^ In previous literature, clinicians have described difficulties providing robotic walking sessions in a clinical setting, such as difficulties with the technology that took up much of the clinical time. Many clinicians believe that robotic walking interventions done in a clinical setting need to occur over longer durations of intervention periods; however, this is challenging in a clinical setting due to constraints within the health care system.^11^ This need for more clinical visits during intervention periods and the travel and time burden of clinical sessions have led to an increased interest in home-based therapies.^**12**^

Novel robotic walking technologies are designed to facilitate home-based therapy. Parents with a child with a disability have stated that they have positive experiences with novel technology intervention.^3–5^ Parents especially enjoyed these interventions because children often experienced an increase in self-confidence and motivation due to an increase in functional improvements. One of these technologies, robotic walkers, facilitate overground walking for children with severe mobility impairments and can be used at home, school, outdoors, and in other areas of a child’s community.^13^ This eliminates barriers families face when trying to access technology in a clinical setting. Robotic walkers can improve physical activity, increase balance, improve sleep and decrease spasticity for children with mobility impairments.^14^ Since this technology can be used at home it, has the potential to create stronger familial bonds. Parents have a strong desire to maximize their child’s therapy and being able to use a robotic walker at home and run the therapy sessions themselves may create more opportunities for children with mobility impairments to engage in therapy.^3,4^

Families’ perspectives of robotic walking as a home-based therapy have yet to be explored, but current home-based therapies have been shown to have many advantages as well as some disadvantages. Parents have commented that home-based therapies are easier to carry out, as they have a low therapy demand for parents, and create satisfaction for both them and their child.^5^ Parents also enjoy being able to see the improvements in their child while they are doing therapies at home.^5^ Parental involvement is crucial for an intervention’s success, as children have shown increased pride and pushed themselves further when their parents were involved in therapy sessions.^4^ As many parents find travelling for therapies to be a major barrier, home-based therapies can be preferable.^15^ Though home-based therapies may be easier for families than clinical therapies, parents have stated that they sometimes find home-based therapies difficult to fit into their daily routine and can cause stress on the caregiver.^3^ Families found home-based therapies particularly difficult when they were a part of a research study and found adhering to the protocol to be hard; therefore, families having a therapy device outside of a study may decrease caregiver stress.^15^ Whether these considerations are relevant for robotic walker use is unknown. An understanding of families’ experiences is necessary to outline the advantages and disadvantages of robotic walkers as a home-based therapy.

## Purpose

The purpose of this study was to explore parents’ perceptions on home-based robotic walking, and how they view the impacts on their family and their child living with a mobility impairment. This study aimed to address the following research questions: What is the experience of families who use a robotic walker in their home and community environment?

## Methods

### Methodology and Design

This study was conducted using a relativist ontology and constructivist epistemology with a collective case study design and thematic analysis method. A relativist ontology assumes that each individual has their own reality, as reality is constructed and created by lived experiences.^16^ It allows us to understand that each family has their own unique experience with robotic walking. A constructivist epistemology understands that knowledge is gained through interacting with others and understanding their unique lived experiences.^17^ An important tenet of this epistemology is that it acknowledges that the researcher themselves influences the research process through their interaction with the data and research participants.^17^ A collective case study design was used because it allows the researchers to examine multiple systems (families) that share a common experience (using a robotic walker). This design allows the researchers to view each family as their own system, therefore allowing for detailed examination of a family’s experience, and then the ability to compare across families.

### Robotic Walker Device

Each family recruited for this study had acquired a Trexo robotic walker (Trexo Robotics, Mississauga, ON). The Trexo robotic walker is designed to be used outside of a clinical setting for children with severe mobility impairments. This device is comprised of two robotic legs, each with motors at the hip and knee and is attached to a Rifton Pacer frame (Rifton Equipment, Rifton, NY). The Trexo is designed for individuals <150lbs who have femur lengths <36cm and calf lengths <44cm. The configuration of this device can be adapted for each individual who uses it. These configurations include using it with or without a saddle and having the participant face posteriorly or anteriorly, with more trunk control needed to face posteriorly. The device is controlled through a tablet interface connected through WIFI that is controlled by a parent or caregiver. During a training session the Trexo records the cadence (steps/minute), “percent initiation” (this variable represents the percent decrease in required forces at each joint only when a joint is moving against gravity and it can be impacted by spasticity and other factors that are not related to the participant initiating movement), time used, and number of steps taken

### Participants

We contacted participants who agreed to participate in a previous study examining self-directed Trexo use. There were 53 parents who were contacted, 10 agreed to participate in the study, but 3 did not respond to interview scheduling emails. As a result, 7 parents participated in this study. All families who participated had been using a Trexo for at least 6 months. The device users (children of parents interviewed) were aged 2-22 (median = 5) years old. There were 4 male and 3 female participants. The majority of the device users had cerebral palsy (6/7), and one had a rare progressive genetic condition.

### Data Collection

Ethics approval for this study was obtained from the University of Calgary Conjoint Health and Research Ethics Board (REB-0799). Six individuals (5 parents and 1 user) from four families with lived experience using the robotic walker reviewed and gave feedback on the interview guide. Each parent who participated was sent a detailed email regarding what the consent form entailed, and they signed the consent form before participating in the interview. Consent forms were also reviewed with the participants before the interview started, so all participants were well informed of what the interview entailed. Interviews were conducted via Zoom (Zoom Communication Inc., USA) and took 21-55 minutes (median = 33 minutes). The interviewer (JLY or AJH) followed a semi-structured focus group guide that asked the participants about their experiences with using the robotic walker, particularly questions related to how it impacts their child using the device and their family as a whole. Each participant was represented by the alphanumeric code used in the parent study.

### Data Analysis

Interviews were audio-recorded and transcribed verbatim and checked by the first author to ensure accuracy of the transcript. NVivo 15 software was used for data management and analysis. Thematic analysis was used to understand and analyze the data in six steps. First, the first author (JLY) read and re-read all the transcripts to ensure accuracy of the transcripts and to gain familiarity with the data. Next, all the transcripts were read through to identify data meaningful to the research question. In this step, the first and last authors (JLY and AJH) both inductively created codes for one transcript to promote coding consistency. The first author then grouped similar codes to create themes and reviewed with co-authors (EGC and AJH). This occurred several times, as themes were reviewed to ensure they represented the lived experiences of the participants. Each theme was created in light of the F-words and Bowen’s family systems theory. The generated themes were checked to ensure they not only represent the lived experiences accurately but also ensure they represent the research question. Theme names were created to express the overarching meaning described by the parents. Following this inductive thematic analysis, themes were deductively categorized into one or more of the F-words. Finally, this report was written to share the results.

### Study Rigour

The rigour for this study was judged against criteria that aligned with the relativist ontology, constructivist epistemology and thematic analysis approach. The first author (JLY) kept a reflectivity journal, where she reflected on the interviews she conducted and any biases that came up during the analysis and writing process. AJH and EGC worked as critical friends throughout the analysis and writing process to help ensure the analysis represented the lived experiences of the participants. Thick description was used by providing rich detail and contextual information so the reader can connect the findings to their own lived experiences.^18^

## Results

The seven interviews conducted resulted in five themes examining how robotic walking impacted family bonding, the physical impacts of using the robotic walker, how robotic walking compares to other therapies, parents’ perspective on the logistics of the device and what it’s like purchasing a robotic walker. These themes were the mapped to four of the F-words, including Family, Fun, Function, and Fitness. Topics related to Friends and Future were not mentioned during the interviews.

### Theme 1: Using a robotic walker at home has led to an increase in family bonding, as families interact with child while robotic walking

Robotic walking has had a significant impact on families and led to family bonding in several ways. Many of these parents had previously only seen their child take a few steps at a time. As a result, they were shocked, excited and proud seeing their child in the robotic walker for the first time. When describing this one mother stated, “I mean, I started crying. There’s so many doctors that have said [device user’s name] will never do this… Getting to see her actually walk was pretty emotional…” (TH-245). This feeling led to the parents feeling pride while seeing their child in the robotic walker. “We were just like so proud of how well she did, given…she had such challenging first experiences with other pieces of equipment that she’s gotten” [TH-186]. It was clear that seeing their child walk was very new and exciting for families, and the parents were happy to have walking as a regular part of their child’s life. When one mom was discussing this, she stated, “…he was out of a wheelchair, and he was moving… It was very powerful” (TH-201). This excitement shows that this technology impacts not only the device user but the family as a whole.

For many families, the robotic walker became a family activity and something the whole family did together. Parents discussed how their whole family gets together and cheers on the child while they are doing their robotic walking session, with on mom stating, “She has a full-blown fan club. [While] [device user’s name] is in the Trexo we’ll just come downstairs and just cheer for her.” (TH-245). Parents also discussed that the whole family would come together and find ways to entertain the child while they were walking. One family discussed how they made their whole basement an obstacle course and would play tag with the child. This is something they have never been able to do as a family before and this has created more fun within the family and how they interact.

Multiple parents also mentioned how the device user’s sibling got involved in the walking sessions and were excited to see their sibling walking. One mom mentioned how the device user’s “…sister, will cheer her on when we’re walking down the street. [Device User’s Name] likes that, or she thinks it’s funny sometimes.” (TH243). Siblings played and did different activities together while in the robotic walker. One dad mentioned how his other children entertain the device user during the sessions, “Sometimes our oldest son will read a book to him… or do his flashcards like his words from school, or they’ll watch a video together” (TH-260). These interactions while the child was in the device made robotic walking fun for the child and changed the families’ view on using therapy equipment. Many of these experiences are new for the family and cheering the child on and interacting with them in new ways had led to an increase in family bonding and stronger family relations.

### Theme 2: Children walk more because they enjoy robotic walking, leading to parents noticing physical changes in child

Many of the parents discussed how their child enjoyed the robotic walker. Many of the device users enjoyed robotic walking, with one mom stating, “She loves it. She loves to be in the Trexo… She loves to be standing and walking and moving” [TH-243]. While many children enjoyed the robotic walker, it is important to mention that one child did not enjoy it and had a negative experience while walking in it for the first time. With their mom stating, “I felt he was going through an anxiety attack” [TH-164].

Since many of the children enjoyed walking, they walked a lot and their parents felt they were getting more physical activity than they typically would. One parent mentioned that their child, “…has a manual chair, and my goal for him was to get more weight bearing, obviously starting with moving his body.” [TH-164]. This statement was echoed by many parents, as an increase in walking and getting more exercise was a major goal for them with the use of the robotic walker. One dad mentioned:

> “Therapy, it just it wasn’t enough. We wanted him to have that motion of his legs moving,… it’s good for him to exercise and not let those muscles get stiff… So that
>
> was the biggest thing, the biggest reason that we pursued it was just we felt you needed that motion of walking for his muscles and exercising” [TH-260].

It was clear that increases in the fitness and function domain were a big motivation for families to seek out and acquire a robotic walker.

Parents were frequently pleased to see an increase in walking and exercise, and this was a common motivation for obtaining a robotic walker. One dad stated how exciting it was “…seeing that he’s at almost 300,000 steps, and seeing that and being like before that, he wasn’t even doing 100 steps in a year, hardly because of just everything” (TH-160). Another parent mentioned how much joy an increase in walking brought her by mentioning, “Most people walk all over the place and that’s all they do. And to see him starting to build that into his life is just very important. Yes, it’s amazing, still amazes me. I’m so happy at the end of each session” (TH-201).

Parents also noticed some physical changes in their child. While discussing how frequent spasticity is an issue for his child, one dad mentioned, “We were hoping it would keep his legs more loose, and it does. It does help with that. Because they’re actually being pulled apart on a daily basis and actually exercised, so that has definitely been good, and that is a huge problem for him” (TH-260). Other physical changes parents noticed were improvements in trunk and head control, an overall increase in strength, and an improvement in bowel movements. When discussing bowel movements, one dad mentioned, “His bowel movements are better, because obviously the exercise makes a difference” (TH-260).

### Theme 3: Parents and children have mixed opinions on robotic walking compared to other therapies

Parents and their children previously struggled with therapies because the sessions were short, and their child wasn’t motivated to participate. Parents felt that their child was getting more repetitions and walking practice while using the robotic walker compared to previous therapies. One mom felt like this increase in repetition was important for neural control of movement. She stated, “ …the more she moves and takes those steps the same way and in the right pattern, and it just like strengthens her body and strengthens her motor pathways” (TH-243). While most parents felt like the robotic walker helped their child more than other therapies, one parent did mention, “For his trunk and stuff like that …I would say horse therapy was better” (TH-186).

Parents also discussed how previous therapies were not as fun for their child, and it was typically difficult to get their child to do other therapy activities. However, for many, using a robotic walker was much more engaging. Seeing their children having fun and engaging more in therapy meant a lot to the parents. When discussing how her child felt in the robotic walker, one mom stated, “She’s just in these devices [other therapy devices], and she’s sitting or standing. But she’s being rolled around or pushed around, whereas the Trexo was so in the moment and engaging for her” (TH-186). Many parents agreed with this sentiment, and one also mentioned that their child used to like other therapies, such as stretching, but no longer enjoys those now that they are using the robotic walker.

One of the main things mentioned by parents was that the burden of travelling and finding specialists for rehabilitation for their child caused parental stress and missed school for their child. Many parents mentioned how they used to travel quite far for physiotherapy appointments. When discussing the impact this had on his family one dad mentioned,

> “We have 2 younger children. And obviously, either I was taking him to therapy, or my wife is taking him to therapy… Sometimes we would drive up to 45 min one way, just to get into therapy, because we don’t have good therapists close here where we’re at and so that has been good not to have to twice a week. Go somewhere, you know. And then he’s getting way more out of school because he’s not being jerked out [for therapy appointments]” (TH-260).

Parents discussed how no they feel much less stress now that they no longer have to go to therapy appointments, which has had an impact on their family. One dad mentioned struggling with finding physiotherapy for his adult child which caused him stress. Having the robotic walker has been beneficial as they now feel like their child’s therapy needs are being covered by using the robotic walker.

### Theme 4: Parents enjoy certain robotic walker features but wish others could be improved

Some parents found the technical aspects of the robotic walker to be quite easy, where others struggled with certain aspects. Specifically related to the control tablet, some parents felt that it was straightforward, whereas others mentioned that they didn’t have much previous technical knowledge and found navigating the tablet to be difficult. When discussing the technological knowledge needed on mom mentioned, “It makes it easier if you have some sort of like just technical strength, makes it easier for using it” (TH186).

There were specific tablet features that the majority of the parents enjoyed. One of these features was being able to see how many steps their child had taken. When discussing the excitement with seeing steps increase, one parent stated, “…he always wants to see what his total number is of steps, you know, sees that climbing up, and he’s excited about that” (TH-260). Another tablet feature parents enjoyed was being able to see an increase in their child’s initiation, with one mom stating, “When she first started, I think she was at like 6 or 7% and she’s consistently just gotten better and better, and we sit there and we watch her, and we’re just everybody’s giving her high fives, and we’re all giving each other high fives to higher her numbers” (TH-245). Checking steps and initiation became a way for the family to bond as they were all excited being able to see clear progress in the device user’s walking.

There were many logistical aspects of the robotic walker that parents discussed. For some parents, getting their child in and out of the robotic walker was very easy. But other parents mentioned that it was difficult. One mom discussed how she was unable to put her child in the robotic walker by herself, “It’s challenging right like cause if she gets upset and she flops backward like, it can be dangerous” (TH-186). One of the major struggles families faced was the logistics of transporting the robotic walker. Parents mentioned that the device is quite big and can be cumbersome to transport. When discussing transporting and using the device outside their home, one parent mentioned, “Another main problem was that it’s really big. And he couldn’t like he couldn’t feasibly use it to go out of the car on the sidewalk into the gym… we’d have to bring his wheelchair, too, and it was really hard to fit everything, we couldn’t fit the Trexo, plus his powerchair for sure. So, we’d have to do the manual chair, and then you have to take the wheels off, and then it’s like shoved in the front seat, so that was hard” (TH-164).

The parents also discussed the accessibility and design of the robotic walker. All the families who were interviewed used the robotic walker at home with a treadmill that can be purchased with it. Parents discussed how they enjoyed the treadmill as it made using the device in their home much more accessible, “We have a house that we bought for with him in mind, so you can go around the main floor. But it was, you know, okay getting coming near the rug you have to lift up the wheel, and then coming near the table you have to move it, you know. So, it was kind of crazy. The treadmill is really nice” (TH-201). One of the main design aspects parents felt needed improvement was the steering of the robotic walker. Currently, the device can only be steered by the parent or caregiver; the device user has no control over steering. Many parents mentioned that this aspect of the device decreases their child’s independence while using the robotic walker. One mom stated, “I was really hoping to find something he could control, and we could go to Target together, and instead of being in a chair, he would be walking. But since he couldn’t steer it, he couldn’t control the speed. He couldn’t do anything independently in it” (TH-164). This sentiment was echoed by many other parents.

### Theme #5: Purchasing the robotic walker was difficult for many families

Robotic walkers are expensive pieces of equipment (current price ∼$50,000 CAD), and families mentioned that acquiring a device was difficult. Families acquired the Trexo in multiple different ways, three families paid for the whole device on their own, one family purchased the Trexo but got the Rifton pacer covered by insurance, two families fundraised for the Trexo, and one family was gifted the Trexo. One mom mentioned how they had to fundraise for the Trexo or else they would not have been able to get a Trexo for their child. She stated, “Cost was high. Insurance was not helping and so it was definitely a struggle at first, because we were like, well, this is what we want to give for her. But we don’t know how to get there. And so we ended up doing a GoFundMe, and we were able to come up with the funds” (TH-245). Beyond the base fee of the Trexo there is also a maintenance package that can be difficult for families to pay for. When discussing this fee one parent said, “Home Care package, the cost was a substantial burden. And we actually cut that off recently” (TH-260).

Since the Trexo is so expensive, it impacted not only how the parents view the company but it also impacted how the families used the Trexo. One parent mentioned that they feel like they should get more support from Trexo based on how much they paid for the device by stating, “*…*that’s been a big pain of just having to do things ourselves” (TH-245). The price of the Trexo also limited how willing families were to use it outside.

**Table 1:**
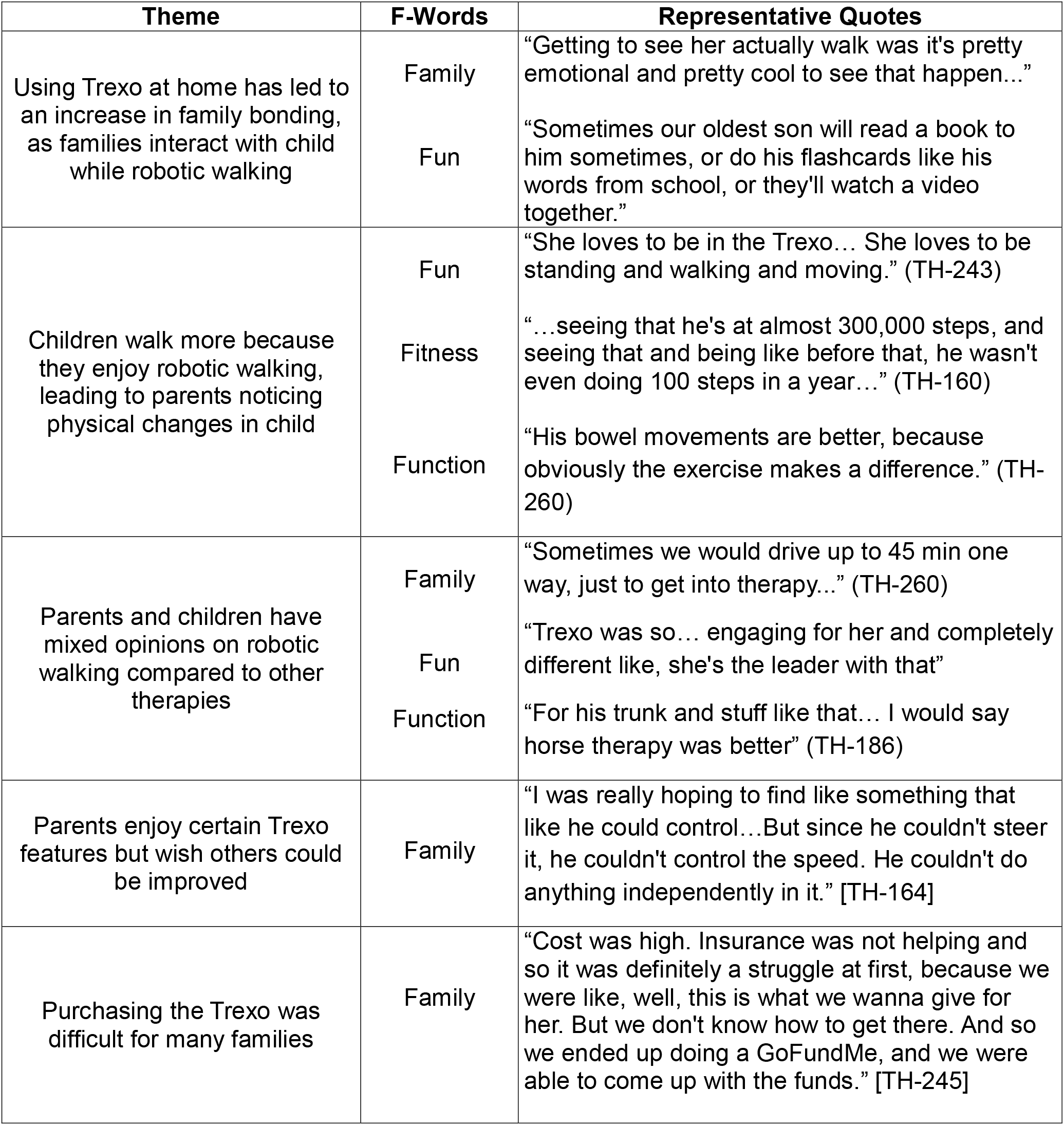
This table shows how each theme relates to the F-words with supporting quotes.

## Discussion

This qualitative study provides insight into families’ experiences using a robotic walker at home. Themes related to the F-words of Family, Fun, Fitness and Function emerged, with parents having both negative and positive experiences within these themes. This deeper understanding families’ experiences can be used to inform family and clinician decisions around whether robotic walkers are an option for home therapy.

Family was the F-word that emerged most frequently across themes, which is not surprising given that the focus of our question was on families. Results emphasize how home-based use of a robotic walker involves Family in ways that clinical-based interventions may not. Parents mentioned an increase in support within the family, which likely resulted from the new approaches to bonding while using the robotic walker and the opportunity to see the child with a disability as more capable, particularly important as it is associated with better psychological and social well-being.^19^

The increase in support and communication across the family may also impact family members and positively increase their emotional functioning. The home-based nature of this intervention enabled a family-centred approach, which is known to lead to more positive physical and psychological outcomes.^20^

Beyond an increase in family bonding, parents also mentioned how having a home-based rehabilitation option decreased their stress. Children enjoyed using the robotic walker more than other therapies, and parents no longer had to travel for therapy appointments. In-person rehabilitation appointments can significantly contribute to parental anxiety, mental strain, physical fatigue and disruption to daily life.^2^ Home rehabilitation allows families to choose convenient times for rehabilitation, instead of their lives and schedule being limited by attending clinical appointments.^3^

However, home-based robotic walker use did not universally decrease parental stress. Difficulties with logistics and finances increased parental stress for some. Parents shared that a lack of knowledge with technology led to increased stress, especially when trying to troubleshoot issues with the robotic walker. A major barrier to parent-delivered rehabilitation is insufficient parental knowledge.^12^ This may have contributed to the lack of confidence and time that were identified as barriers in this study. Parents described feeling nervous to use the robotic walker in certain scenarios and struggling to find time to use the Trexo consistently.^12^ These barriers could be addressed by increasing the amount of training parents receive and by supporting a plan of how the robotic walker will fit into their lives before deploying the device. The F-words of Family and Fun were closely intertwined. Home-based use of the robotic walker created opportunities for fun interactions for the device user with their parents and siblings. Parents described how the ways they interacted while their child was using the robotic walker led to both experiencing more Fun than they had with previous therapeutic devices and interventions. Families were excited for robotic walking sessions, encouraging and cheering on their child. This aligns with previous research of children and parents feeling “happier and lighter”, being able to do therapies at home.^3^ Uniquely, the siblings of the device user were also highly engaged in sessions. Typically developing siblings are eager and willing playmates, but they do not often have the opportunity to participate in their siblings’ rehabilitation activities.^21^ Supporting the fun experienced between siblings during rehabilitation sessions is important, as sibling involvement can contribute to positive therapy outcomes.^22^

Parents also described many positive changes in the Fitness and Function domains. It is known that 90% of children with mobility impairments spend most of their days sedentary, which leads to chronic health conditions.^23^ Likely as a result of this, one of the main motivations for using the robotic walker was to increase their child’s physical activity levels. Robotic walking can facilitate light physical activity for individuals with mobility impairments,^24^ which may have led to the physical improvements described by parents. One parent specifically felt it was important to facilitate strengthening of the motor pathways and a recent review found robotic walking aids can indeed facilitate neuroplasticity. While it is beyond the scope of this qualitative study to explore the mechanism underlying physical changes, parents described changes in function, such as increases in strength, trunk control, bowel movements, head control and decreases in spasticity. Previous interventions examining robotic walkers have shown similar results.^14,24,25^

The F-word domain Friends and Future were not discussed by participants in this study. It is possible that since all the families used the robotic walker predominantly at home, children did not have the opportunity to interact with peers while using the robotic walker. Constraining use to the home may have limited children’s social experiences during device use. In a previous study where families used a robotic walker in their community, positive changes in social relationships were clearly noticed by many of the families.^24^ Therefore, using a robotic outside of the home may have additional benefits for children within the Friends domain. Future, was also not asked about within the questions therefore we did not get a clear understanding regarding how robotic walking may impact parents views on their child’s future.

## Limitations

Limitations of this study include factors impacting the generalizability. The results of this study may not be representative of the experiences of all families who have a robotic walker due to the sample size. Also, only one device was used in this study. As a result, the results of this study may not be able to be extrapolated to other robotic walkers designed for children with limited independent walking and home therapies. We chose to conduct these interviews at one time point, once families had a device for at least six months. Future qualitative research would benefit from investigating the ongoing experiences of families at multiple time points.

## Conclusions

This study adds to the limited body of research examining how home-based therapies may impact families and children living with mobility impairments by showing how such therapies impact different aspects of a child’s life. Using a robotic walker at home led to an increase in family bonding, new and fun ways for families and siblings to interact, and improvements in function for children living with a disability. Having a rehabilitation device at home decreased parental stress in general, though some families experienced difficulties with the logistics of using the robotic walker at home. These findings highlight opportunities to support families using robotic walkers as a home-based rehabilitation tool for their child living with a mobility impairment.

## Declaration of Interests

Trexo Robotics provided in-kind support for this study including support for participant recruitment, but academic independence was protected as part of our Data Sharing Agreement. They have not participated in this manuscript’s preparation. Our research team has also received an unrestricted donation from Trexo Robotics ($40,000 in 2023). Dr. Condliffe is an unpaid member of the Clinical Advisory Board at Bionic Power Inc. The authors have no further conflicts of interest to disclose.

## Data Availability statement

The participants of this study did not give written consent for their data to be shared publicly, so due to the sensitive nature of the research, supporting data is not available.

## Notes

### Funding Statement

NSERC Brain CREATE provided funding for this study.

### Author Declarations

The University of Calgary Conjoint Health and Research Ethics Board gave ethical approval for this work (REB-0799).

## References

1. Ono Ericka, Friedlander R, Salih Tamara. BCMJ_Vol61_No3-service-gaps.

2. Fritz HL. Coping with caregiving: Humor styles and health outcomes among parents of children with disabilities. Res Dev Disabil. 2020;104. doi:10.1016/j.ridd.2020.103700

3. Holmes C, Shields N, Morgan P, Brock K, McKenzie G, Reddihough D. Home-based motorised cycling in Non-ambulant adults with cerebral palsy: a feasibility study. Disabil Rehabil. Published online 2024. doi:10.1080/09638288.2024.2353234

4. Phelan SK, Gibson BE, Wright FV. What is it like to walk with the help of a robot? Childrens perspectives on robotic gait training technology. Disabil Rehabil. 2015;37(24):2272–2281. doi:10.3109/09638288.2015.1019648

5. Beckers LWME, Geijen MME, Kleijnen J, et al. Feasibility and effectiveness of home-based therapy programmes for children with cerebral palsy: A systematic review. BMJ Open. BMJ Publishing Group. 2020;10(10). doi:10.1136/bmjopen-2019-035454

6. Rosenbaum P, Gorter JW. The “F-words” in childhood disability: I swear this is how we should think. Child Care Health Dev. 2012;38(4):457–463. doi:10.1111/j.1365-2214.2011.01338.x

7. Stucki G. International classification of functioning, disability, and health (ICF): A promising framework and classification for rehabilitation medicine. In: American Journal of Physical Medicine and Rehabilitation. Vol 84. 2005:733–740. doi:10.1097/01.phm.0000179521.70639.83

8. Soper AK, Cross A, Rosenbaum P, Gorter JW. Exploring the international uptake of the “F-words in childhood disability”: A citation analysis. Child Care Health Dev. Blackwell Publishing Ltd. 2019;45(4):473–490. doi:10.1111/cch.12680

9. McCormick A, Alazem H, Biddiss E, et al. The explotion of technologies in pediatric rehabilitation a call to use the F-Words lens. In: Hayre CM, Muller D, Scherer M, Hackett PMW, Gordley-Smith A, eds. Emerging Technologies in Healthcare. CRC Press; 2024:147–160.

10. Trivette CM, Dunst CJ, Hamby DW. Influences of Family-Systems Intervention Practices on Parent-Child Interactions and Child Development. Topics Early Child Spec Educ. 2010;30(1):3–19. doi:10.1177/0271121410364250

11. Ehrlich-Jones L, Crown DS, Kinnett-Hopkins D, et al. Clinician Perceptions of Robotic Exoskeletons for Locomotor Training After Spinal Cord Injury: A Qualitative Approach. Arch Phys Med Rehabil. 2021;102(2):203–215. doi:10.1016/j.apmr.2020.08.024

12. Massey J, Harniess P, Chinn D, Robert G. Barriers and facilitators to parent-delivered interventions for children with or infants at risk of cerebral palsy. An integrative review informed by behaviour change theory. Disabil Rehabil.Taylor and Francis Ltd. 2025;47(2):287–301. doi:10.1080/09638288.2024.2338193

13. Maggu M, Udasi R, Nikitina D. Designing Exoskeletons for Children: Overcoming Challenge Associated with Weight-bearing and Risk of Injury. ACM/IEEE International Conference on Human-Robot Interaction. 2018;94(8):39. doi:10.1145/3173386.3177840

14. Hilderley AJ, Diot CM, Shen H, et al. Overground robotic walker use in the home and community: a six-month prospective cohort study. J Neuroeng Rehabil. Published online December 13, 2025. doi:10.1186/s12984-025-01812-8

15. Hao J, Huang B, Remis A, He Z. The application of virtual reality to home-based rehabilitation for children and adolescents with cerebral palsy: A systematic review and meta-analysis. Physiother Theory Pract.Taylor and Francis Ltd. 2024;40(7):1588–1608. doi:10.1080/09593985.2023.2184220

16. Ivankova N V, Creswell JW. 7 Mixed Methods.

17. Lincoln YS, Guba EG. PARADIGMATIC CONTROVERSIES/CONTRADICTIONS/AND EMERGING CONFLUENCES. In: 1994.

18. Burke S. Rethinking “validity” and “trustworthiness” in qualitative inquiry: how might we judge the quality of qualitative research in sport and exercise science? In: Routledge Handbook of Qualitative Research in Sport and Exercise. Taylor and Francis, New York, NY; 2016:330–339.

19. King G, Willoughby C, Specht JA, Brown E. Social support processes and the adaptation of individuals with chronic disabilities. Qual Health Res. 2006;16(7):902–925. doi:10.1177/1049732306289920

20. Matt R. Citation: Matt R (2024) Family Centered Approach to Pediatric Physical Therapy: How Involving Parents and Caregivers Enhances Therapeutic Outcomes and Supports Child Development. J Physiother Rehabi. 2024;8(4):4. doi:10.4172/JPTR.1000173

21. Madzimbe P, Maart S, Corten L, Dambi J. Participation of fathers and siblings in home rehabilitation programmes for children with neuro-developmental delay: a scoping review. BMC Pediatr. 2024;24(1). doi:10.1186/s12887-024-05119-w

22. Zagacki LM, Chiarello LA, Palisano RJ, Lieberman-Betz RG. Sibling Participation in Occupational Therapy for Children with Physical Disabilities: A Case Report. Disabilities. 2025;5(3). doi:10.3390/disabilities5030079

23. Martin JJ. Benefits and barriers to physical activity for individuals with disabilities: A social-relational model of disability perspective. Disabil Rehabil. 2013;35(24):2030–2037. doi:10.3109/09638288.2013.802377

24. Youngblood JL, Norman BM, Diot CM, et al. RObotic WAlking for children who CAnnot WAlk (RoWaCaWa): Feasibility and family impacts and perspectives of a family-led Intervention. Preprint posted online December 27, 2025. doi:10.64898/2025.12.19.25341914

25. Diot CM, Youngblood JL, Friesen AH, et al. Robot-Assisted Gait Training with Trexo Home: Users, Usage and Initial Impacts. Children. 2023;10(3). doi:10.3390/children10030437

